# A precision health approach to medication management in neurodevelopmental conditions: a model development and validation study using four international cohorts

**DOI:** 10.1101/2025.03.12.25323683

**Authors:** Marlee M. Vandewouw, Kamran Niroomand, Harshit Bokadia, Sophia Lenz, Jesiqua Rapley, Alfredo Arias, Jennifer Crosbie, Elisabetta Trinari, Elizabeth Kelley, Robert Nicolson, Russell J. Schachar, Paul D. Arnold, Alana Iaboni, Jason P. Lerch, Melanie Penner, Danielle Baribeau, Evdokia Anagnostou, Azadeh Kushki

## Abstract

Psychotropic medications are commonly prescribed to children with neurodevelopmental conditions, but responses vary widely, prescribing is largely off-label, and the expertise required is concentrated in specialized programs. We developed artificial intelligence models to predict prescribing patterns of stimulants, anti-depressants, and anti-psychotics. Feasibility was established in research cohorts by predicting cross-sectional medication use from the Child Behaviour Checklist, with training and internal testing in the Province of Ontario Neurodevelopmental network (*N*=598) and external testing in the Healthy Brain Network (*N*=1,764) and Adolescent Brain Cognitive Development (*N*=2,396) studies. Clinical evaluation used electronic medical records (EMRs) from the Psychopharmacology Program (*N*=312) at Holland Bloorview Kids Rehabilitation Hospital to predict the medication class prescribed at a follow-up visit (∼3 months later). In all cohorts, the modelled outcome was the clinician’s prescribing decision, which reflects clinician judgement, family preference, tolerability, and access to care in addition to expected effectiveness, and does not directly measure treatment response or clinical benefit. In the research cohorts, internal testing achieved an area under the receiver operating characteristic curve (median [IQR]) of 0.75 [0.73,0.80] for stimulants, 0.83 [0.78,0.87] for anti-depressants, and 0.79 [0.72,0.86] for anti-psychotics, and external testing confirmed generalizability. In the EMR cohort, values were 0.84 [0.81,0.88] for stimulants, 0.82 [0.77,0.87] for anti-depressants, and 0.87 [0.83,0.91] for anti-psychotics. Findings demonstrate that AI can accurately learn expert prescribing patterns and predict medication prescribing decisions, supporting the potential of data-driven tools to guide personalized medication management for neurodevelopmental conditions.

## Background

Neurodevelopmental conditions, such as autism spectrum disorder (autism), intellectual disability (ID), and attention-deficit/hyperactivity disorder (ADHD), are highly heterogeneous and overlapping in their biology and phenotypic presentation^1,2^, which includes hundreds of associated genes^3^, and significant variability in neurobiological correlates^2^. Despite decades of research, no markers exist to indicate which biological pathway may be implicated for an individual child to guide individualized care^4,5^, and current approaches to care provision remain reliant on broad diagnostic labels. This lack of precision is particularly challenging for pharmacological interventions in neurodevelopmental conditions. These interventions are highly prevalent with greater than 50% of children with neurodevelopmental conditions prescribed psychotropic medications to address behavioural and/or mental health concerns (e.g., anxiety, hyperactivity, aggressive or self-injurious behavior), and high rates of polypharmacy during childhood (∼30%)^6^.

Despite the prevalent use of psychotropic medications in neurodevelopmental conditions, there is tremendous variability in individual-level responses to psychotropic treatment for co-occurring mental health and behavioural conditions^7^, reflecting the heterogeneity of biological pathways impacted, diverse clinical presentations, as well as sociodemographic complexities that influence the acceptability and uptake of medications. This variability poses a significant challenge in determining the appropriate medication for an individual child, especially in cases where co-occurring conditions (e.g., autism, ADHD, anxiety) can interact and increase the complexity of presentation and response. Efforts to identify biological predictors of treatment response have included magnetic resonance spectroscopy measures of GABA and glutamate to assess responsiveness to glutamatergic and GABAergic agents^8^, electroencephalographic measures of stimulant response^9^, and heart rate variability as a response predictor to adrenergic agents^10^. These approaches remain investigational, however, and require equipment and expertise that are not routinely available in settings where prescribing decisions are made. Prescribing practices therefore remain almost entirely off-label^6,11–15^ and based on trial-and-error approaches that can expose children to multiple medication trials with the potential for severe and life-long adverse effects for some classes of medication^16^. Navigating these decisions therefore relies on the accumulated judgement of experienced clinicians, expertise that is concentrated in specialized programs and unavailable in most community settings where these children receive care.

To fill this gap, we propose an artificial intelligence (AI) model that uses an individual’s characteristics to predict the medication class an experienced clinician would prescribe, among the commonly prescribed classes in neurodevelopmental conditions: stimulants, anti-depressants, and anti-psychotics. We first establish the feasibility of this approach by building models to predict medication usage using a research cohort of children and youth with neurodevelopmental conditions, using two international research cohorts for external validation. Then, to examine how this approach generalizes to real-world clinical settings, we leveraged a patient cohort to predict the medication class prescribed at a follow-up visit approximately 3 months later from electronic medical record (EMR) data of children and youth with neurodevelopmental conditions who visited a specialized psychopharmacology clinic.

## Methods

### Study design and cohorts

For the research cohorts, training and internal testing was conducted using cross-sectional data from 598 children and youth with neurodevelopmental conditions extracted from the Province of Ontario Neurodevelopmental (POND) network (Ontario, Canada). External testing was performed using two cross-sectional international cohorts: 1,764 children and youth with neurodevelopmental conditions from the Healthy Brain Network (HBN^17^; New York, United States), and 2,396 children and youth with neurodevelopmental conditions from the Adolescent Brain Cognitive Development study (ABCD^18^; United States).

For clinical validation, longitudinal EMRs from the Psychopharmacology Program (PPP) at Holland Bloorview Kids Rehabilitation Hospital (Toronto, Ontario, Canada) were used. PPP sees children with neurodevelopmental conditions who have tried at least one psychoactive medication without adequate success. Data were extracted from electronic medical records of 312 children who visited the PPP over a three-year period (2019 – 2022) and returned for a follow-up approximately 3 months later.

Participant demographics are summarized in Table 1. Further details on the cohorts, including eligibility and consent procedures, and a comparison of the research cohorts can be found in the Supplementary Material. This study was conducted in accordance with the Declaration of Helsinki, and was approved by the Research Ethics Board at Holland Bloorview Kids Rehabilitation Hospital (#1854).

**Table 1.** Cohort demographics and medication outcomes.

|  |  | Research cohorts |  |  | EMR cohort |
| --- | --- | --- | --- | --- | --- |
|  |  | POND | HBN | ABCD | PPP |
| *N* |  | 598 | 1,764 | 2,396 | 312 |
| Age range (years) |  | 5 – 19 | 5 – 17 | 8 – 11 | 4 – 17 |
| Mean age [SD] (years) |  | 11.4 [3.2] | 10.1 [2.7] | 9.9 [0.6] | 10.4 [2.9] |
| Sex [%] | Male | 424 [71] | 1,213 [69] | 1,523 [64] | 262 [84] |
|  | Female | 174 [29] | 551 [31] | 873 [36] | 50 [16] |
| Autism [%] |  | 338 [57] | 325 [18] | 0 [0] | 292 [94] |
| ADHD [%] |  | 193 [32] | 1,381 [78] | 1,751 [73] | 181 [58] |
| OCD [%] |  | 67 [11] | 90 [5] | 1,048 [44] | 16 [5] |
| Intellectual disability [%] | Yes | 84 [14] | 86 [5] | 149 [6] | 105 [34] |
|  | No | 514 [86] | 1,678 [95] | 2,247 [94] | 135 [43] |
|  | Unknown | 0 [0] | 0 [0] | 0 [0] | 72 [23] |
| Race/ethnicity [%] | White | 327 [55] | 871 [49] | 1,257 [52] | NA |
|  | Minoritized | 156 [26] | 866 [49] | 1,137 [47] | NA |
|  | Not collected | 115 [19] | 27 [2] | 2 [0] | 312 [100] |
| Annual household income [%] | Low | 86 [14] | 307 [17] | 770 [32] | NA |
|  | Medium | 98 [16] | 345 [20] | 608 [25] | NA |
|  | High | 170 [28] | 786 [45] | 798 [33] | NA |
|  | Not collected | 244 [41] | 326 [18] | 220 [9] | 312 [100] |
| Outcome [%] | Stimulant | 171 [29] | 177 [10] | 551 [23] | 114 [37] |
|  | Anti-depressant | 96 [16] | 72 [4] | 142 [6] | 82 [26] |
|  | Anti-psychotic | 52 [9] | 26 [1] | 43 [2] | 123 [39] |

EMR: electronic medical record; PPP: Psychopharmacology Program; POND: Province of Ontario Neurodevelopmental network; HBN: Healthy Brain Network; ABCD: Adolescent Brain Cognitive Development study; SD: standard deviation; ADHD: attention-deficit/hyperactivity disorder; OCD: obsessive-compulsive disorder. For POND, primary diagnosis is reported; for ABCD, HBN, and PPP, refer to supplemental.

### Predictors and outcome

For the longitudinal PPP cohort, the set of predictors was co-created with the clinicians at the PPP program to reflect features relevant to the medication classes considered, and the availability of variables in the electronic medical records at the first visit. These predictors were broadly grouped into five categories: demographics, clinical, family-related, child-related, and care-related, the last of which included prior use of psychotropic medication classes at the first visit. Further detail on the predictors is presented in the Supplementary Material. The outcome was whether a medication of a given class was prescribed at the follow-up visit, approximately three months after the first visit, for each of three primary classes: stimulants, anti-depressants, and anti-psychotics (Table 1). Non-stimulants were considered in a supplementary analysis, given variability in the clinical classification of this medication group.

For the cross-sectional research cohorts, demographics (age and sex), neurodevelopmental diagnoses (autism, ADHD, and OCD), IQ as a continuous score, and data from the Child Behaviour Checklist (CBCL)^19^ were used as predictors. Further detail on the predictors is provided in the Supplementary Material, including a between-cohort comparison of the demographics and CBCL scores (Supplemental Tables 1 and 2). The outcome was whether a child was taking a medication of a given class (stimulants, anti-depressants, anti-psychotics) at the time of study (Table 1).

**Table 2.**
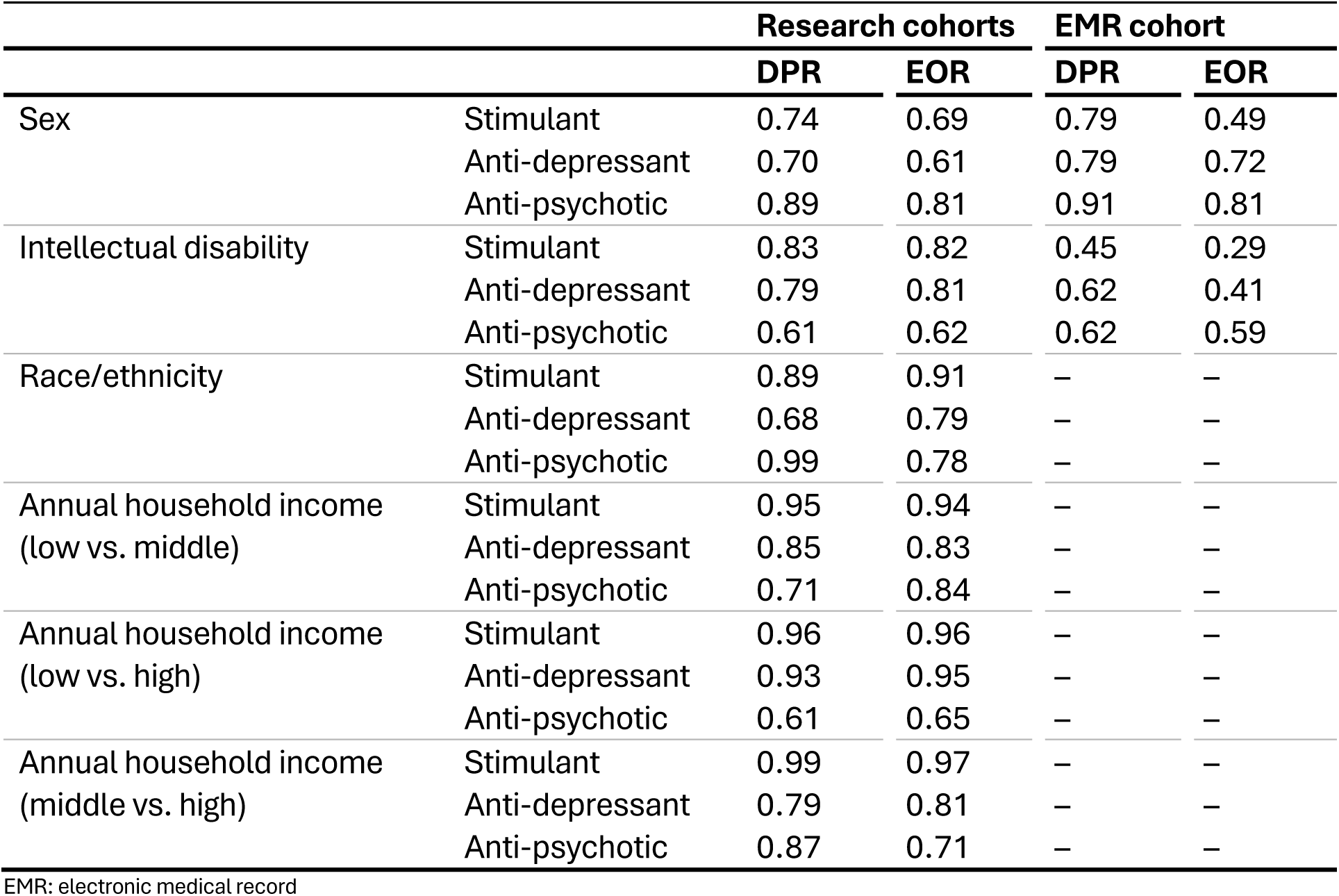
Demographic parity ratio (DPR) and equalized odds ratio (EOR) for sensitive groups.

|  |  | Research cohorts |  | EMR cohort |  |
| --- | --- | --- | --- | --- | --- |
|  |  | DPR | EOR | DPR | EOR |
| Sex | Stimulant | 0.74 | 0.69 | 0.79 | 0.49 |
|  | Anti-depressant | 0.70 | 0.61 | 0.79 | 0.72 |
|  | Anti-psychotic | 0.89 | 0.81 | 0.91 | 0.81 |
| Intellectual disability | Stimulant | 0.83 | 0.82 | 0.45 | 0.29 |
|  | Anti-depressant | 0.79 | 0.81 | 0.62 | 0.41 |
|  | Anti-psychotic | 0.61 | 0.62 | 0.62 | 0.59 |
| Race/ethnicity | Stimulant | 0.89 | 0.91 | – | – |
|  | Anti-depressant | 0.68 | 0.79 | – | – |
|  | Anti-psychotic | 0.99 | 0.78 | – | – |
| Annual household income<br>(low vs. middle) | Stimulant | 0.95 | 0.94 | – | – |
|  | Anti-depressant | 0.85 | 0.83 | – | – |
|  | Anti-psychotic | 0.71 | 0.84 | – | – |
| Annual household income<br>(low vs. high) | Stimulant | 0.96 | 0.96 | – | – |
|  | Anti-depressant | 0.93 | 0.95 | – | – |
|  | Anti-psychotic | 0.61 | 0.65 | – | – |
| Annual household income<br>(middle vs. high) | Stimulant | 0.99 | 0.97 | – | – |
|  | Anti-depressant | 0.79 | 0.81 | – | – |
|  | Anti-psychotic | 0.87 | 0.71 | – | – |

EMR: electronic medical record

### Model

AI models were built to predict the outcome from the set of predictors, separately for each medication class (Figure 1). For the research cohorts, the models were trained and internally tested using the POND dataset, while HBN and ABCD were withheld for external testing. By validating with two external datasets, we assessed the generalizability and robustness of the models, ensuring results are not specific to a single dataset and can be replicated across diverse populations. For the EMR cohort (PPP), only internal testing was performed.

**Figure 1.**
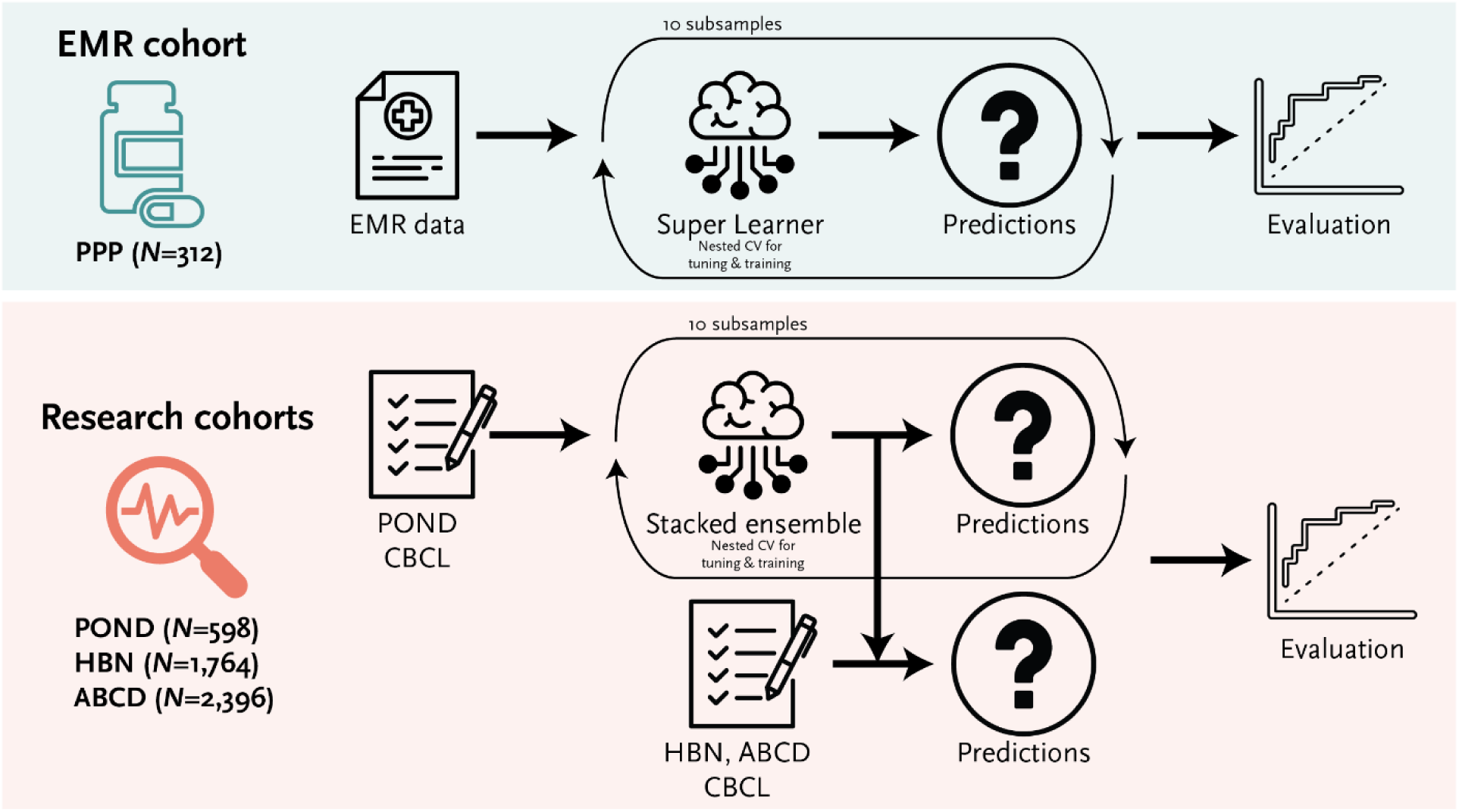
Study design.

To address class imbalance, we applied random under-sampling of the majority class to achieve approximately balanced 1:1 class ratios. This algorithm-agnostic approach reduces majority-class dominance and strengthens the minority-class learning signal without relying on synthetic data or class weighting. To limit dependence on any single balanced draw, we repeated the under-sampling for 10 independent iterations, noting that findings were stable across 5, 10, and 20 iterations.

For the AI models, we implemented a Super Learner approach^20^, combining CatBoost gradient boosting^21^ and deep neural networks as base learners, with logistic regression meta-learner trained on out-of-fold predictions to generate final predictions. We used nested cross-validation with inner 3-fold loops for hyperparameter optimization using Optuna^22^ and outer 5-fold loops where base learners generated out-of-fold predictions for the logistic regression meta-learner. Further details are provided in the Supplementary material, including a comparison of the Super Learner with its individual base learners and with logistic regression.

### Performance evaluation

Model performance was assessed using the area under the receiver operating characteristic curve (AU-ROC), reporting medians and interquartile ranges (IQR) across folds and under-sampling iterations. Additional metrics (area under the precision-recall curve (AU-PR), sensitivity, and specificity), and model calibration (calibration slope, calibration intercept, and Brier score) are provided in the Supplementary Material.

To interpret which features were most important for predicting longitudinal medication choice, Shapley Additive exPlanations (SHAP) values^23^ were computed for the EMR (PPP) models. SHAP values were computed for each of the base models and combined by taking their weighted average using the coefficients from the logistic regression meta-learner.

The demographic parity and equalized odds ratios^24,25^ were used to evaluate whether the model predictions were independent of sensitive group status for sex, intellectual disability (ID), race/ethnicity, and annual household income. The metric was computed at the default probability threshold (0.50) across folds, under-sampling iterations, and datasets for the research cohorts, where predictions from the five cross-validation models were averaged for each participant within each under-sampling iteration before pooling across iterations and cohorts, so that each participant contributes equally regardless of whether evaluation was internal (POND) or external (HBN, ABCD). The 80% rule was used to determine any biases^26^.

## Results

### Model performance

The research cohorts demonstrated the feasibility of AI predicting treatment status cross-sectionally for each medication class (Figure 2), with internal testing (POND) demonstrating good-to-excellent AU-ROC that was significantly higher than chance for stimulants (median [IQR]: 0.75 [0.73, 0.80]), anti-depressants (0.83 [0.78, 0.87]), and anti-psychotics (0.79 [0.72, 0.86]). The performance of the external testing sets (HBN and ABCD) indicated good generalizability, with the AU-ROCs slightly reduced but still in the good-to-excellent range. Additional metrics (area under the precision-recall curve, sensitivity, specificity) are reported in Supplemental Table 3, model calibration in Supplemental Figure 1 and Supplemental Table 4, and a comparison with the individual base learners and logistic regression in Supplemental Table 5. For the EMR cohort (PPP; Figure 3A), the AU-ROC across the cross-validation folds was excellent for stimulants (0.84 [0.81, 0.88]), anti-depressants (0.82 [0.77, 0.87]), and anti-psychotics (0.87 [0.83, 0.91]). Predictive performance for non-stimulants is reported in Supplemental Table 6.

**Figure 2.**
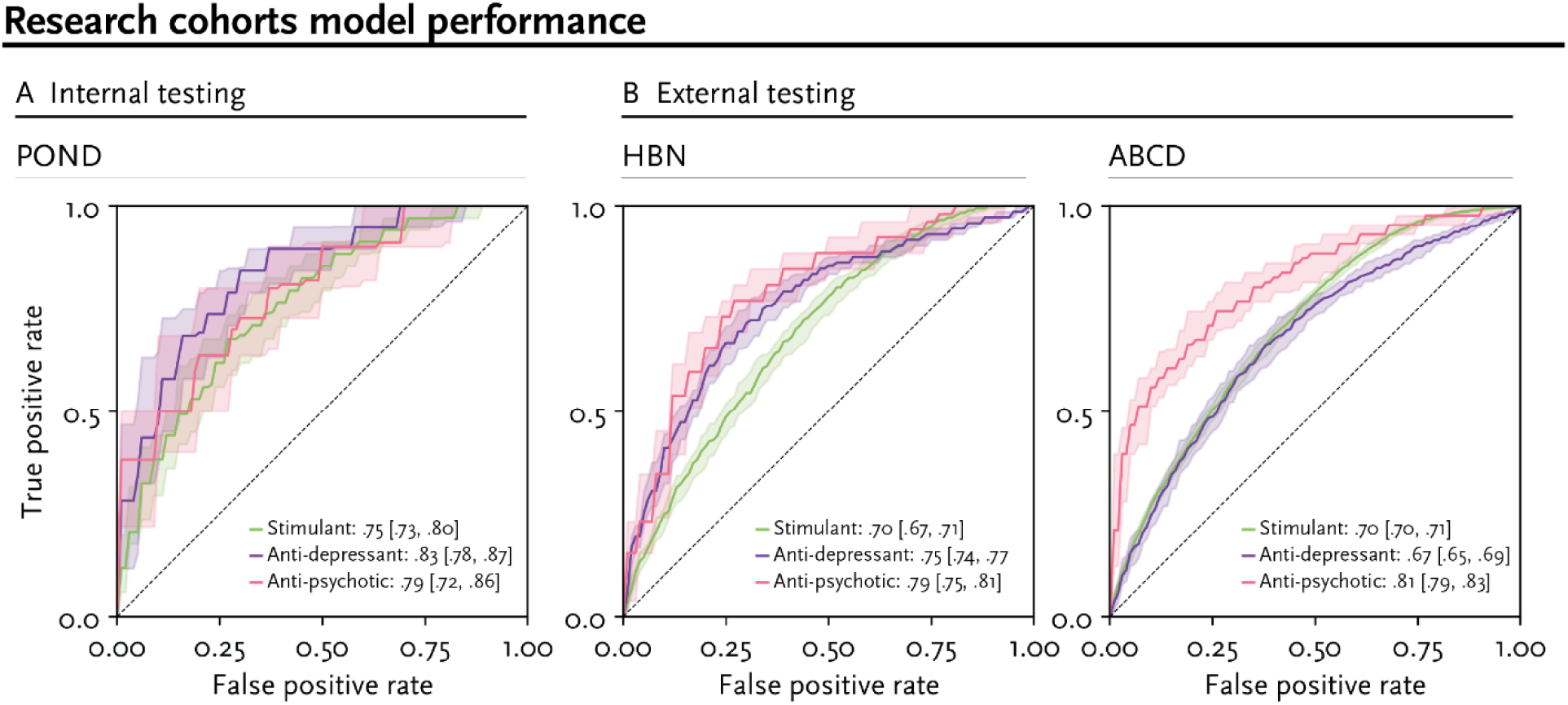
ROC curves for the research cohorts, for the internal testing (A; POND) and external testing (B; HBN and ABCD) sets for each of the medication classes (green: stimulants, purple: anti-depressants, pink: anti-psychotics); AU-ROCs are also presented. POND: Province of Ontario Neurodevelopmental network; HBN: Healthy Brain Network; ABCD: Adolescent Brain Cognitive Development study; ROC: receiver operating characteristic curve; AU-ROC: area under the ROC

**Figure 3.**
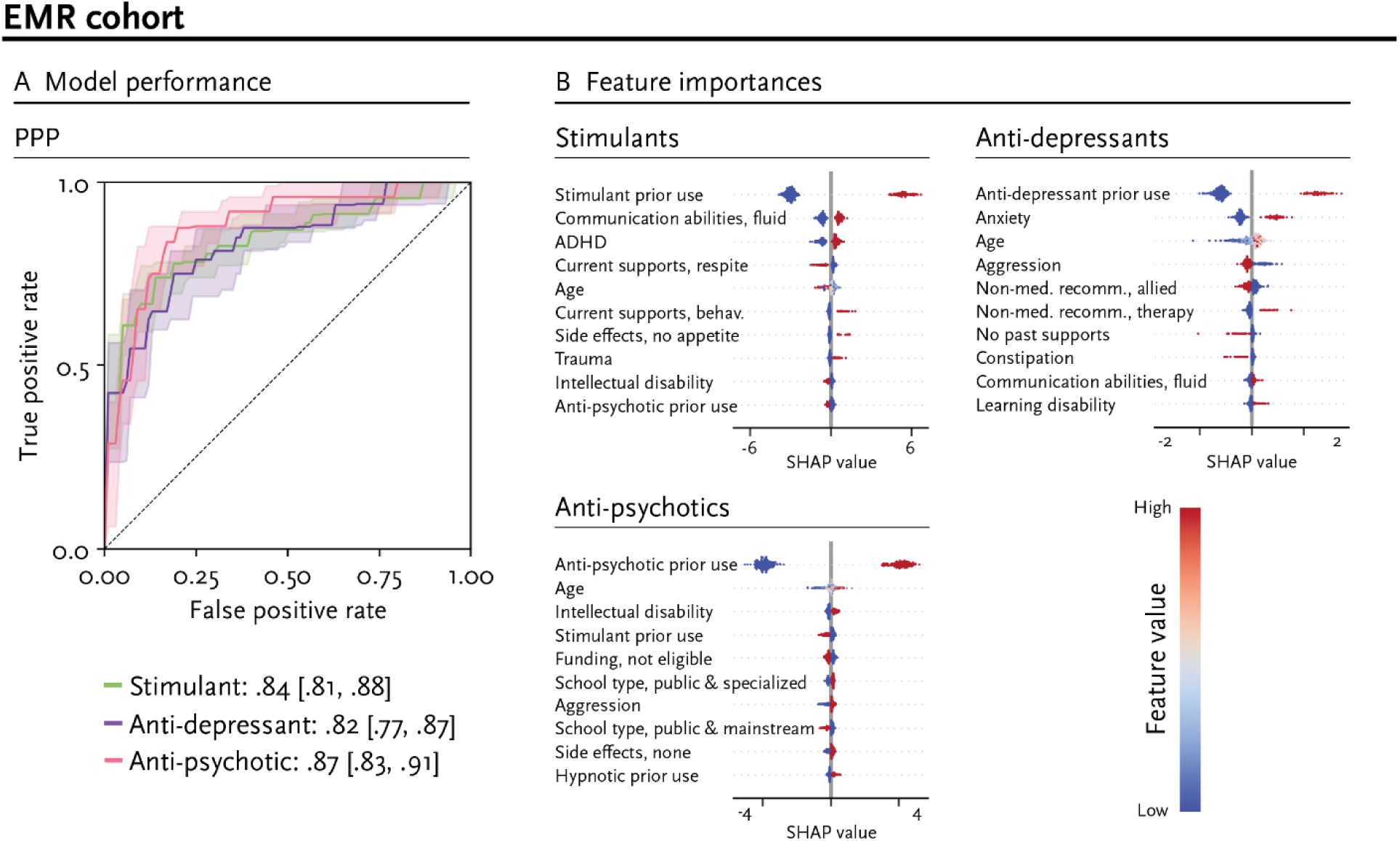
(A) ROC curves for the EMR cohort (PPP) for each of the medication classes (green: stimulants, purple: anti-depressants, pink: anti-psychotics); AU-ROCs are also presented. (B) Feature importance for the EMR cohort (PPP) assessed using SHAP values. The SHAP values are plotted for each participant for the 10 most important features for each medication class, sorted by importance (mean absolute SHAP value) from top to bottom. Colours represent the feature value (red for high values, blue for low values), indicating whether higher or lower values contribute positively or negatively to prediction scores. EMR: electronic medical record; PPP: Psychopharmacology Program; ROC: receiver operating characteristic curve; AU-ROC: area under the ROC; SHAP: Shapley Additive exPlanations; ADHD: attention-deficit/hyperactivity disorder

### Feature importance

SHAP values were computed for the EMR cohort (PPP), to identify which features were most important for predicting medication choice (Figure 3B). Prior use of a medication class was the most important feature across all three classes. In particular, prior use of a medication within the same class, increased the likelihood of a medication choice in that class. Age was also a salient feature: younger, middle, and older participants were more likely to be recommended stimulants, anti-psychotics, and anti-depressants, respectively. Other important features included ADHD and communication abilities for stimulants, anxiety and aggression for anti-depressants, and intellectual disability and prior prescription of stimulants for anti-psychotics. In a sensitivity analysis, re-fitting of the EMR model after removing the prior medication-use features reduced AU-ROCs as expected, but all models remained significantly above chance (Supplemental Table 7), indicating that the remaining predictors contribute independent signal.

### Bias

The demographic parity and equalized odds ratios were computed to evaluate the bias of the models (Table 2), where values below 0.80 indicate bias. To support interpretation of the ratios, per-group metrics are provided in Supplemental Tables 8 and 9 for the research and EMR cohorts, respectively. For the research cohorts, parity was mixed. Individuals who were male were more likely to be recommended a stimulant (female, male: 46%, 62%). Individuals who were female, did not have an ID, who were white, and who were of high income class were more likely to be recommended an anti-depressant (female, male: 48%, 33%; ID, no-ID: 31%, 39%; white, minority: 43%, 29%; low, middle, high income: 38%, 32%, 41%). On the other hand, those with an ID, and those with a lower annual household income were more likely to be recommended anti-psychotics (ID, no-ID: 68%, 41%; low, middle, high income: 56%, 40%, 34%). In the EMR cohort, males were more likely to be recommended stimulants (male, female: 46%, 36%), females were more likely to be recommended anti-depressants (male, female: 44%, 55%), and children with an ID were more likely to be recommended anti-psychotics (ID, no-ID: 64%, 39%) and less likely to be recommended stimulants (ID, no-ID: 28%, 61%). This was in line with trends in the data.

To determine whether observed disparities reflect or amplify existing prescribing patterns, we compared the model-predicted DPR to baseline DPR computed from actual prescribing labels (Supplemental Table 10). In the research cohorts, the model largely tracked baseline prescribing. The model moved DPR above the threshold for anti-psychotic predictions by sex, where baseline prescribing was disparate (baseline DPR=0.72, model DPR=0.89). Conversely, the model introduced a meaningful disparity in anti-depressant predictions for sex (baseline DPR=0.93, model DPR=0.70). In the EMR cohort, no attribute meaningfully crossed the four-fifths threshold in either direction.

## Discussion

Medication management for children with neurodevelopmental conditions remains a significant challenge, particularly in community settings where subspecialty expertise may not be available. To address this gap, this study aimed to develop and evaluate an AI algorithm to predict clinician prescribing decisions in neurodevelopmental conditions. Ultimately, this type of tool could be used as a decision-aid to support medication management where specialized expertise is not available.

We showed that AI can learn expert clinician medication decisions with high accuracy. We also demonstrated that the results generalize across independent, international datasets, which is notable given the drastically different characteristics of these cohorts, in terms of demographics, diagnosis, phenotype, and region-specific access to service and supports (see Supplemental Tables 1 and 2). While the accuracy in these research cohorts is lower compared to the EMR model, this is likely due to the cross-sectional nature of the prediction in these cohorts. For example, use of stimulant medications to target ADHD behaviours confounds the measures of ADHD traits when considered cross-sectionally, in turn impacting predictive performance.

Overall, these results support the feasibility of using an AI-based decision aid tool. This can be particularly advantageous for community practitioners, where subspecialty expertise required for medication management is often not available. These findings show that the prescribing decisions of an experienced clinician can be learned from routinely collected data and reproduced across independent cohorts; they do not show that prescribing guided by such a model would improve symptom outcomes, tolerability, or the number of medication trials a child undergoes. These are separate claims – the value proposition in community settings is the transfer of specialist prescribing patterns to contexts where that expertise is unavailable, whereas whether this also improves clinical outcomes remains to be established. This transfer is necessarily bounded by the expert clinical judgement being reproduced, which is itself imperfect, but the alternative in these settings is prescribing that is largely off-label and proceeds by trial-and-error in the absence of clinical guidelines. Prospective trials are needed to further validate this type of algorithm in clinical settings.

Explainability and interpretability are critical features of responsible AI systems, contributing to enhancing trust and transparency with both clinicians and patients^27^. To promote explainability, we characterized the relative contributions of clinical and demographic features to the AI algorithm’s medication recommendations. For the EMR cohort, these contributions were closely aligned with indications for each medication class (e.g., stimulants for ADHD, SSRIs for anxiety). Contributing features also reflected sociodemographic and environmental factors that may contribute to clinical decision making beyond primary indications. For example, school environment and funding eligibility were significant contributors to the decision to recommend an anti-psychotic. These findings highlight the complexity of the clinical and sociodemographic factors that contribute to value-laden decisions around medication management for children with neurodevelopmental conditions.

The strongest contributor for all three classes was prior use of a medication in that class. This finding reflects the fact that medication choices often remain within the same class of medication but entail changes in the medication type and dosing within that class (e.g., amphetamines, methylphenidate within the stimulant class). Future work will focus on adding this type of resolution to the AI predictions. Although prior use was the strongest contributor, a sensitivity analysis re-fitting the model without any of the prior use features showed that performance decreased but remained significantly above chance across all three classes. This is expected given that PPP, by design, serves children who have already tried one or more psychoactive medications – making prior exposure a core input to every clinical decision at this program – but also indicates that the remaining demographic, clinical, family-related, child-related, and care-related features carry independent predictive signal beyond current medication status. Because prior exposure is inherently associated with subsequent prescribing, however, continuity of care and individualized treatment response cannot be fully separated in observational prescribing data. These characteristics of our dataset also mean our results speak to medication optimization rather than first-line selection, for which established clinical guidelines already exist.

The features used here are those routinely captured in clinical settings, which makes such models deployable where care is delivered and, as our findings indicate, captures contextual factors such as school environment and funding eligibility that no biological measures would reflect. Nonetheless, many of the features used describe observable presentation rather than the biological mechanisms underlying treatment response, and biological markers^8–10^ may eventually provide complementary information in this respect, though they are not yet validated for this purpose or available in routine care. The framework used here is agnostic to predictor type, so such measures could be incorporated and their incremental value assessed as they become available.

Another aspect of responsible AI is potential algorithmic biases that may disproportionately disadvantage subgroups of individuals. Our analysis revealed several biases including increased recommendation of anti-psychotics to children who had intellectual disability and were from lower income households. On the other hand, anti-depressants recommendations were increased for children who did not have an intellectual disability, and who were white. It is important to note that these biases are not related to any specific feature of the AI algorithm, but largely mirror previously reported biases in prescription patterns which exist across the system^28^. For example, decreased access to early diagnosis and behavioural interventions for children from lower income household may contribute to a higher likelihood of crisis situations that necessitate prescription of anti-psychotic medications. Similarly, it is well established that depression can be challenging to detect in individuals with an intellectual disability^29^.

Comparison of model-predicted to baseline prescribing disparities showed that the model largely reproduced existing prescribing patterns. In the research cohorts, the model attenuated the disparity in anti-psychotic prescribing by sex, but introduced a new disparity in anti-depressant predictions by sex that was not present in the underlying prescribing data. This may partly reflect sex differences in presentation on behavioural measures such as the CBCL. No attribute showed amplified disparity in the EMR cohort, which most closely reflects the intended clinical population for real-world implementation. This is particularly relevant for community settings – the intended deployment context for this type of tool – which often serve children from more marginalized backgrounds. Because the model is trained on historical prescribing data, there is a risk that it could reproduce existing inequities; our baseline comparison indicates that it largely does so, without substantially amplifying them. Computational bias mitigation strategies, such as fairness constraints during model training and group-specific calibration, can be applied to further reduce predicted disparities, though these technical approaches alone cannot resolve the broader system-level inequities that drive disparate prescribing patterns.

### Limitations

The results of this study should be interpreted considering several limitations. Firstly, for the longitudinal prediction, we defined outcome as the medication choice at the follow-up visit. Treatment decisions in neurodevelopmental conditions are a complex, subjective, and value-laden construct. The use of medication choice was co-created with clinicians as an outcome as it reflects the aggregate effects of the multitude of factors contributing to a clinical decision to not stop a medication (e.g., ineffectiveness, side-effects, acceptability or other factors). These children also visited the specialized clinic after at least one psychoactive medication was tried without success; our findings therefore speak to medication optimization rather than first-line selection, which would require evaluation in a cohort that is medication-naïve at baseline. Prescribing practice in community settings, the ultimate deployment setting for this type of tool, also differs from that of a specialized program, and the model’s utility in this context remains to be established. The research cohorts were also cross-sectional, with predictors measured while children were taking the medications being predicted; associations in these models should therefore not be interpreted directionally. Although gender identity and race/ethnicity were not included as predictors due to incomplete capture in the primary EMR cohort, their omission may contribute to model misspecification bias. These variables remain important contextual factors for understanding medication access and prescribing patterns. Future work should incorporate reliably captured measures and evaluate model performance across gender and racial/ethnic groups, alongside fairness assessments, updating the model as needed to mitigate potential inequities. The models were trained using data exclusively from North America; future work should investigate their translation to other global populations to account for potential differences in healthcare systems, prescribing practices, and sociocultural influences on medication usage. Finally, this study exclusively focused on predicting medication classes; these classes are broad, and future work should examine within-class predictability.

## Conclusions

In summary, the present study focused on AI model development and evaluation, demonstrating that clinician prescribing decisions for children with neurodevelopmental conditions – which capture factors including observed treatment response, tolerability, family preference, and access to care – can be predicted from routinely collected data. To enable effective clinical implementation of this type of AI-tool, future studies are needed to evaluate the algorithm prospectively at the point of care, including its effect on clinical outcomes, its acceptability, and the barriers and facilitators to uptake.

## Supplementary material

### PPP

#### Dataset description

The clinical cohort used for this study was longitudinal electronic medical records from the Psychopharmacology Program (PPP) at Holland Bloorview Kids Rehabilitation Hospital (Toronto, Canada). The PPP is a program with provincial catchment in Ontario that provides medication management for children and youth with neurodevelopmental conditions. Children were eligible for this study if (1) their first visit was between 2019-2022, (2) they were at least four years of age at their first visit, and (3) they returned to the clinic for a follow-up visit. As the data was extracted from electronic medical records, no consent was obtained. Data from PPP is not publicly available due to ethics restrictions. Clinical data from all longitudinal clinical visits were extracted into a RedCap database from semi-structured medical record templates by trained chart reviewers. Two chart reviewers co-reviewed each medical record in sets of five and reviewed discrepancies until 85% agreement on all items was achieved. Reviewers proceeded independently after that, and any uncertainties were discussed with the study team.

#### Predictors

For PPP, the predictors were grouped into five categories: demographics, clinical, family-related, child-related, and care-related. Categorical variables with unknown or undocumented values were dummy-coded with ‘Unknown’ as the implicit reference category. This retains all participants and prevents missing data from being conflated with a substantive level. Missingness ranged between 0 and 28% across predictors, and 65% of participants had at least one missing predictor.

Demographic variables included age, sex (male or female), and region (urban (Toronto, Greater Toronto Area, or other Ontario city) or rural (rural or northern community)). Due to missing data, gender and race/ethnicity could not be included as predictors. To capture socioeconomic status, whether a parent was receiving social assistance or means-tested disability support as a source of income was included as a predictor.

Clinical variables included neurodevelopmental diagnoses (attention-deficit/hyperactivity disorder (ADHD), autism, obsessive-compulsive disorder (OCD), intellectual disability, learning disorder, other psychiatric condition), mental health symptoms (anxiety, depression, irritability), and chronic conditions (epilepsy, cerebral palsy, asthma, migraines, gastroesophageal reflux disease, persisting constipation, obesity, and other).

Family-related factors included a family history of psychiatric or developmental diagnoses in a first degree relative, and family reports of conflict between family members in the household, caregiver burnout, and financial barriers to care, as reported by families and documented by the treating clinician in the child’s chart.

Child-related factors included school type (public, mainstream classroom, private, mainstream classroom, public, specialized classroom, private, specialized classroom, homeschool, or other), school attendance (full days, part days or withdrawal, frequent calls home for pick-up, frequent school refusal or frequent absences, not attending or demitted from school, or other), communication abilities (speaks fluidly in sentences, speaks a few words or short phrases, non-speaking and uses communication technique, non-speaking and no other formal communication form, and other). History of trauma (children’s aid involvement, witnessing violence, experiencing violence, physical abuse, sexual abuse, or neglect, other significant stressor as perceived by the child, or other stressor), police involvement, hospitalization, emergency department visits, intensive crisis service involvement due to behavioural or mental health concerns, aggression, self-injury, and suicidal thoughts were also included.

Care-related factors included receiving government funding for care (specifically, the Ontario Assistance for Children with Severe Disabilities Program), which was coded as “yes”, “has not applied”, “not eligible”, or “other”. Data on current, past, and recommended supports were collected using the following categories: (1) behavioural therapy, Applied Behaviour Analysis (ABA), or parent coaching by a behaviour therapist, (2) child therapy (e.g., Cognitive Behavioural Therapy (CBT) or group therapy), (3) case management, (4) allied health, (5) respite, (6) other, or (7) none. Prior use of five commonly prescribed medication classes (stimulants, non-stimulants, hypnotics, anti-depressants, and anti-psychotics) was determined as whether a child was taking a medication at the time of the first visit to the clinic. The behaviour that was targeted for treatment at the first visit was collected using the following categories: physical aggression, swearing/verbal threats, elopement, self-injury, verbal refusal, repetitive behaviours, property destruction, tantrums/meltdowns, and/or other. Data on side effects reported at the first visit were categorized as: headache, stomach-ache, constipation, nausea/vomiting, weight gain, weight loss, irritability, drowsy/fatigue/sedation, insomnia, appetite decrease, appetite increase, other, and/or none. Whether adherence problems for medications were discussed at the first visit was also included as a predictor.

#### Outcome

The predicted outcome was the medication class the child was on at the follow-up visit.

#### Bias

Bias was assessed on sex (male versus female) and intellectual disability (yes versus no).

### POND

#### Dataset description

The POND network is a research network spanning several institutions across Ontario, Canada that collects demographic, phenotypic, and biological data from children and youth with and without neurodevelopmental conditions. Participants were eligible for the current study if they received a diagnosis of autism, ADHD, or OCD, if they were between 5 and 19 years of age, and if diagnostic, medication usage, IQ, and the Child Behaviour Checklist (CBCL^19^) was available. Children and youth provided written informed consent if they were able; otherwise, they provided verbal assent, and their caregiver provided written consent. Data from POND is managed by the Ontario Brain Institute and is available through a controlled data release. All requests for access should be directed to the Brain-CODE platform (https://www.braincode.ca/content/controlled-data-releases#dr006).

#### Predictors

Predictors for POND included demographics (age and sex), diagnosis (autism, ADHD, or OCD), IQ, and the CBCL. Diagnoses were confirmed with expert clinical judgement alongside the Autism Diagnostic Observation Schedule, 2 (ADOS-2^30^) and Autism Diagnostic Interview, Revised (ADI-R^31^) for autism, the Parent Interview for Child Symptoms (PICS^32^) and/or the Kiddie Schedule for Affective Disorders and Schizophrenia (K-SADS^33^) for ADHD, and the Children’s Yale-Brown Obsessive-Compulsive Scale (YBOCS^34^) for OCD. Full-scale IQ was collected using age-appropriate Wechsler^35–39^ or Stanford-Binet^40^ scales. All the individual questions, syndromic scale *T*-scores (anxious/depressed, withdrawn/depressed, somatic complaints, social problems, thought problems, attention problems, rule-breaking behaviour, and aggressive behaviour), and summary *T*-scores (internalizing problems, externalizing problems, and total problems) from the CBCL were included as predictors.

#### Outcome

Caregivers were asked to provide a list of medication their child was taking. These medications were classified as stimulants, non-stimulants, anti-depressants, and anti-psychotics with assistance from clinicians. The predicted outcome was the medication class(es) the child was on.

#### Bias

Bias was assessed for sex, intellectual disability, race/ethnicity, and socioeconomic status. For sex, biological sex (male versus female) was used. Intellectual disability (yes versus no) was determined using the full-scale IQ cutoff of 70. For race/ethnicity, participants/caregivers were asked to select all groups that applied: Aboriginal, Arab, Black, Chinese, East Asian, Filipino, Japanese, Korean, Latin American/Hispanic, South Asian, Southeast Asian, West Asian, White, other; this data was used to categorize individuals as white or minoritized. For socioeconomic status, we used annual household income categorized as low (<$50,000), middle ($50,000-$99,999), and high (≥$100,000).

### HBN

#### Dataset description

HBN is a mental health study of children and adolescents across the New York city (United States) area that has collected demographic, phenotypic, and biological data; families who had psychiatric concerns about their child were recruited for the study. Participants were eligible for the current study if they received a diagnosis of autism, ADHD, or OCD, if they were between 5 and 19 years of age, and if diagnostic, medication usage, IQ, and the CBCL^19^ was available. Participants at least 18 years of age provided written informed consent; otherwise, they provided written assent, and their caregiver provided written consent. Access to the data used in this study is protected by a Data Usage Agreement (DUA; see https://fcon_1000.projects.nitrc.org/indi/cmi_healthy_brain_network for more details), which was awarded to AK for this study.

#### Predictors

Aligning with POND, predictors for HBN included demographics (age and sex), diagnosis (autism, ADHD, and/or OCD), full-scale IQ, and the CBCL. For diagnosis, the consensus DSM-5^41^ diagnoses provided by the HBN clinical team were used. Full-scale IQ was assessed using Wechsler scales^38,39^ or the Kaufman Brief Intelligence Test^42^.

#### Outcome

Caregivers were asked to provide a list of medication their child had taken the day of the visit. These medications were classified as stimulants, non-stimulants, anti-depressants, and anti-psychotics with assistance from clinicians. The predicted outcome was the medication class(es) the child was on.

#### Bias

Bias was assessed on sex, intellectual disability, race/ethnicity, and socioeconomic status. For sex, biological sex (male versus female) was used. Intellectual disability (yes versus no) was determined using the full-scale IQ cutoff of 70. For race/ethnicity, participants/caregivers indicated their ethnicity (not Hispanic or Latino, Hispanic or Latino, decline to specify, or unknown) and race (White/Caucasian, Black/African American, Hispanic, Asian, Indian, Native American Indian, American Indian/Alaskan Native, Native Hawaiian/Other Pacific Islander, two or more races, or other race), which were used to categorize individuals as white or minoritized. For socioeconomic status, we used annual household income categorized as low (<$50,000), middle ($50,000-$99,999), and high (≥$100,000).

### ABCD

#### Dataset description

The ABCD study^43^ is a longitudinal study of adolescents from the general population across the United States that has collected demographic, phenotypic, and biological data. For the purposes of this study, only the first time point (collected at approximately 9-11 years of age) was used. Participants were eligible for the current study if medication usage, IQ, the Kiddie Schedule for Affective Disorders and Schizophrenia (K-SADS), and the CBCL was collected. Caregivers provided written informed consent, and participants provided written assent. Access to the data used in this study is protected by an NIMH Data Archive DUA for the Adolescent Brain Cognitive Development (ABCD) permission group (see https://nda.nih.gov/user/dashboard/data_permissions.html for more details), which was awarded under DUA #15619 for this study.

#### Predictors

Aligning with POND, predictors for ABCD included demographics (age and sex), diagnosis (autism, ADHD, and/or OCD), IQ, and the CBCL. Diagnoses were assigned using a computerized version of the K-SADS^44^ completed by parents and/or youth. IQ was assessed using the NIH Toolbox^45^ Cognition Battery’s age-corrected total cognition composite score, which shows a high degree of alignment with the Wechsler scales used in POND^46^.

#### Outcome

Caregivers were asked to provide a list of medication their child had taken in the two weeks preceding the visit. These medications were classified as stimulants, non-stimulants, anti-depressants, and anti-psychotics with assistance from clinicians. The predicted outcome was the medication class(es) the child was on.

#### Bias

Bias was assessed on sex, intellectual disability, race/ethnicity, and socioeconomic status. For sex, biological sex (male versus female) was used. Intellectual disability (yes versus no) was determined using the IQ cutoff of 70. For race/ethnicity, participants/caregivers indicated their race/ethnicity (Hispanic/Latino, non-Hispanic Asian, non-Hispanic Black, non-Hispanic White, and Multi-Racial/Other), which were used to categorize individuals as white or minoritized. For socioeconomic status, we used annual household income categorized as low (<$50,000), middle ($50,000-$99,999), and high (≥$100,000).

#### AI models

We implemented a Super Learner approach^20^ in Python (version 3.12), combining CatBoost gradient boosting^21^ (version 1.2.7) and deep neural networks (Keras^47^ (version 3.7.0) with TensorFlow^48^ (version 2.18.0)) as base learners, with a logistic regression model trained on out-of-fold predictions to generate final predictions.

Prior to model training, continuous predictors were standardized to zero mean and unit variance, with the transformation fit on the training data of each cross-validation fold and applied to the corresponding validation fold and external testing data.

Hyperparameters for both base learners were optimized using Optuna’s Tree-structured Parzen Estimator^22^ with 100 trials and nested 3-fold cross-validation within each training fold, with both models optimizing AU-ROC. The deep neural network architecture was tuned across 1-3 hidden layers, 48-96 units per layer, and dropout rates of 0.20-0.40. Networks used ReLU activation and the Adam optimizer, with learning rates tuned between 3×10^-4^ to 3×10^-3^. Training used a fixed batch size of 32 with early stopping (patience=12) monitoring AU-ROC over a maximum of 200 epochs. CatBoost models were configured with 50-400 iterations, learning rates of 0.01-0.30, tree depths of 3-6, and early stopping after 200 rounds without improvement.

The Super Learner consisted of a logistic regression model (L2 penalty = 1.0, liblinear solver) to combine predicted probabilities from both base learners. We employed nested cross-validation, with inner 3-fold loops for hyperparameter optimization and outer 5-fold loops in which base learners trained on 4 folds generated out-of-fold predictions for the logistic regression model. This design followed standard Super Learner methodology to prevent overfitting, avoid data leakage, and yield unbiased performance estimates.

#### Comparison of research cohorts

The demographic variables (age, sex, diagnosis, intellectual disability, race/ethnicity, and socioeconomic status) were compared amongst the research cohorts (Supplemental Table 1). Chi-squared tests were used for categorical variables, holding significance at *p*<.050 and reporting Cramer’s *v* effect sizes. Shapiro-Wilks tests indicated that all continuous variables were non-normally distributed; as such, Kruskal-Wallis tests were used, holding significance at *p*<.050, and reporting eta-squared effect sizes.

**Supplemental Table 1.** Demographic comparison of the research cohorts.

| Measure | *p*-value | Effect size | Post-hocs |
| --- | --- | --- | --- |
| Age | <.0001 | .05 | ABCD > POND > HBN |
| Sex | .0001 | .06 | Females: ABCD > POND, HBN |
| Autism | <.0001 | .53 | POND > HBN > ABCD |
| ADHD | <.0001 | .32 | HBN > ABCD > POND |
| OCD | <.0001 | .43 | ABCD > POND > HBN |
| Intellectual disability | <.0001 | .11 | POND > HBN, ABCD |
| Race/ethnicity | <.0001 | .10 | White: POND > HBN, ABCD |
| Annual household income | <.0001 | .13 | HBN > POND > ABCD |

POND: Province of Ontario Neurodevelopmental network; HBN: Healthy Brain Network; ABCD: Adolescent Brain Cognitive Development study; ADHD: attention-deficit/hyperactivity disorder; OCD: obsessive-compulsive disorder

The CBCL summary *T*-scores were also compared amongst the datasets (Supplemental Table 2), using Kruskal-Wallis tests due to non-normality of all variables (Shapiro-Wilks tests), holding significance at *p*<.050, and reporting eta-squared effect sizes.

**Supplemental Table 2.**
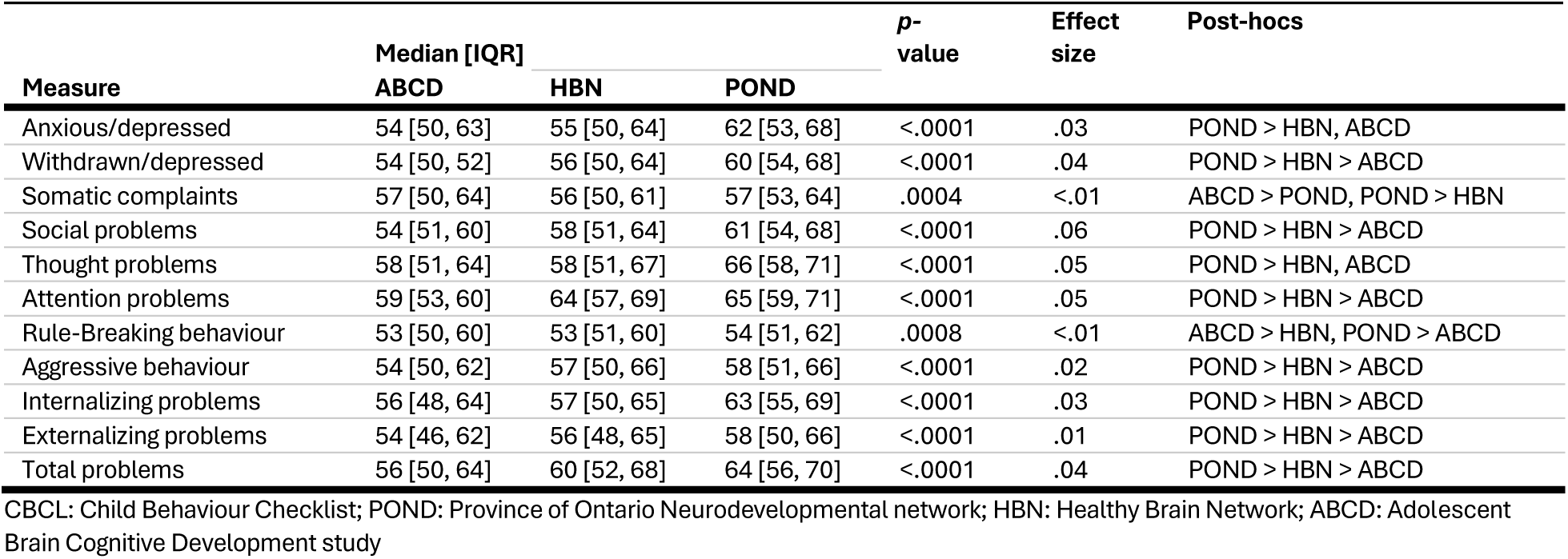
CBCL comparison of the research cohorts.

| Measure | Median [IQR] |  |  | *p*-value | Effect size | Post-hocs |
| --- | --- | --- | --- | --- | --- | --- |
|  | ABCD | HBN | POND |  |  |  |
| Anxious/depressed | 54 [50, 63] | 55 [50, 64] | 62 [53, 68] | <.0001 | .03 | POND > HBN, ABCD |
| Withdrawn/depressed | 54 [50, 52] | 56 [50, 64] | 60 [54, 68] | <.0001 | .04 | POND > HBN > ABCD |
| Somatic complaints | 57 [50, 64] | 56 [50, 61] | 57 [53, 64] | .0004 | <.01 | ABCD > POND, POND > HBN |
| Social problems | 54 [51, 60] | 58 [51, 64] | 61 [54, 68] | <.0001 | .06 | POND > HBN > ABCD |
| Thought problems | 58 [51, 64] | 58 [51, 67] | 66 [58, 71] | <.0001 | .05 | POND > HBN, ABCD |
| Attention problems | 59 [53, 60] | 64 [57, 69] | 65 [59, 71] | <.0001 | .05 | POND > HBN > ABCD |
| Rule-Breaking behaviour | 53 [50, 60] | 53 [51, 60] | 54 [51, 62] | .0008 | <.01 | ABCD > HBN, POND > ABCD |
| Aggressive behaviour | 54 [50, 62] | 57 [50, 66] | 58 [51, 66] | <.0001 | .02 | POND > HBN > ABCD |
| Internalizing problems | 56 [48, 64] | 57 [50, 65] | 63 [55, 69] | <.0001 | .03 | POND > HBN > ABCD |
| Externalizing problems | 54 [46, 62] | 56 [48, 65] | 58 [50, 66] | <.0001 | .01 | POND > HBN > ABCD |
| Total problems | 56 [50, 64] | 60 [52, 68] | 64 [56, 70] | <.0001 | .04 | POND > HBN > ABCD |

CBCL: Child Behaviour Checklist; POND: Province of Ontario Neurodevelopmental network; HBN: Healthy Brain Network; ABCD: Adolescent Brain Cognitive Development study

#### Performance evaluation

Additional metrics are provided in the Supplemental Table 3, including the area under the precision–recall curve (AU-PR; where recall = sensitivity and precision = positive predictive value [PPV]), along with sensitivity and specificity.

**Supplemental Table 3.** Model performance.

|  | Research cohorts |  |  | Clinical cohort |
| --- | --- | --- | --- | --- |
|  | Internal testing | External testing |  | Internal testing |
|  | POND | HBN | ABCD | PPP |
| **Median AU-PR [IQR]** |  |  |  |  |
| Stimulant | .76 [.72, .80] | .66 [.65, .69] | .67 [.66, .69] | .87 [.84, .90] |
| Anti-depressant | .83 [.77, .89] | .75 [.73, .77] | .66 [.63, .68] | .86 [.80, .90] |
| Anti-psychotic | .83 [.75, .88] | .78 [.73, .81] | .83 [.81, .86] | .86 [.81, .90] |
| **Median sensitivity [IQR]** |  |  |  |  |
| Stimulant | .71 [.65, .77] | .77 [.72, .81] | .67 [.63, .73] | .74 [.66, .78] |
| Anti-depressant | .79 [.68, .84] | .49 [.47, .54] | .42 [.36, .49] | .71 [.56, .88] |
| Anti-psychotic | .71 [.61, .80] | .69 [.54, .77] | .58 [.51, .67] | .88 [.79, .88] |
| **Median specificity [IQR]** |  |  |  |  |
| Stimulant | .68 [.60, .74] | .50 [.45, .55] | .60 [.55, .65] | .87 [.78, .91] |
| Anti-depressant | .74 [.68, .80] | .84 [.82, .86] | .80 [.75, .84] | .81 [.69, .88] |
| Anti-psychotic | .70 [.50, .80] | .79 [.69, .85] | .88 [.79, .93] | .84 [.79, .88] |

PPP: Psychopharmacology Program; POND: Province of Ontario Neurodevelopmental network; HBN: Healthy Brain Network; ABCD: Adolescent Brain Cognitive Development study; AU-PR: area under the precision recall curve; IQR: interquartile range

#### Calibration

Calibration of the models was assessed using the calibration slope, the calibration intercept (calibration-in-the-large, estimated with the slope fixed at one), and the Brier score, computed from the predicted probabilities pooled across folds, and under-sampling iterations (Supplemental Table 4). Calibration curves were constructed by dividing predictions into ten equal-sized bins and plotting the observed proportion against the mean predicted probability within each bin (Supplemental Figure 1). As models were trained and evaluated on class-balanced samples, predicted probabilities are calibrated to a 50% event rate; Brier scores are therefore interpreted relative to the value of 0.25 expected from an uninformative model on balanced data. Calibration-in-the-large was close to ideal in the internal testing sets, while deviations from zero were observed for some medication classes in the external testing sets. Calibration slopes exceeded one for most models, indicating that predicted probabilities were conservative, tending toward the middle of the range rather than the extremes.

**Supplemental Table 4.** Model calibration.

| Attribute | Outcome | Research cohorts |  |  | EMR cohort |
| --- | --- | --- | --- | --- | --- |
|  |  | Internal testing<br>POND | External testing<br>HBN | ABCD | Internal testing<br>PPP |
| Slope | Stimulant | 1.23 | 0.98 | 0.99 | 1.42 |
|  | Anti-depressant | 1.40 | 1.05 | 0.71 | 1.67 |
|  | Anti-psychotic | 1.70 | 1.79 | 2.03 | 1.34 |
| Intercept | Stimulant | -0.03 | -0.24 | -0.05 | -0.06 |
|  | Anti-depressant | -0.06 | 0.42 | 0.40 | -0.06 |
|  | Anti-psychotic | -0.07 | 0.08 | 0.15 | -0.06 |
| Brier score | Stimulant | 0.20 | 0.23 | 0.22 | 0.16 |
|  | Anti-depressant | 0.18 | 0.21 | 0.24 | 0.18 |
|  | Anti-psychotic | 0.21 | 0.21 | 0.20 | 0.14 |

EMR: electronic medical record; PPP: Psychopharmacology Program; POND: Province of Ontario Neurodevelopmental network; HBN: Healthy Brain Network; ABCD: Adolescent Brain Cognitive Development study

**Supplemental Figure 1.**
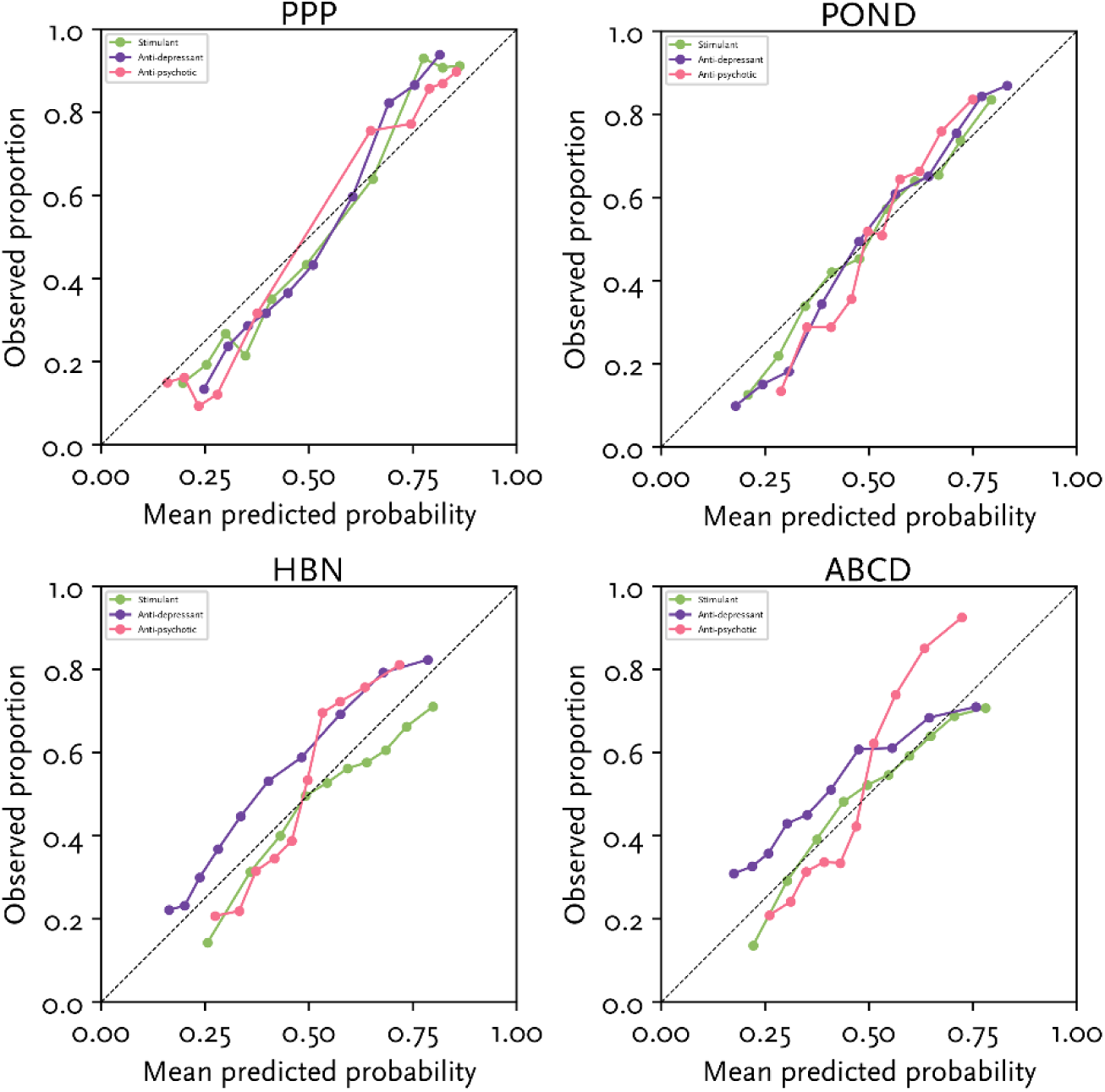
Calibration curves.

#### Model comparison

To assess whether the Super Learner improved on its constituent models, we compared it with each base learner evaluated alone (CatBoost and the deep neural network) and with logistic regression as a conventional comparator (Supplemental Table 5). The Super Learner outperformed logistic regression in every cohort and medication class, by 0.02 to 0.08 AU-ROC. Differences from the individual base learners were smaller – the Super Learner performed as well or better than both base learners in 8 of 12 cohort-class combinations and was within 0.01 AU-ROC of the better base learner in the remainder. Neither base learner was consistently superior, with CatBoost performing better in the EMR cohort with predominantly categorical predictors, and the neural network performing better for anti-psychotic prediction in POND and ABCD. The ensemble therefore provided performance close to the best available base learner in every setting without requiring the better model to be identified in advance.

**Supplemental Table 5.** Comparison of the Super Learner with its base learners and a conventional model.

| Cohort | Outcome | Super Learner | CatBoost | Neural network | Logistic regression |
| --- | --- | --- | --- | --- | --- |
| POND | Stimulant | 0.75<br>[0.73,0.80] | 0.75<br>[0.71,0.78] | 0.75<br>[0.72,0.79] | 0.67<br>[0.63,0.70] |
|  | Anti-depressant | 0.83<br>[0.78,0.87] | 0.81<br>[0.76,0.86] | 0.81<br>[0.77,0.86] | 0.75<br>[0.68,0.81] |
|  | Anti-psychotic | 0.79<br>[0.72,0.86] | 0.75<br>[0.70,0.81] | 0.80<br>[0.72,0.85] | 0.74<br>[0.68,0.81] |
| HBN | Stimulant | 0.70<br>[0.67,0.71] | 0.69<br>[0.68,0.71] | 0.67<br>[0.64,0.69] | 0.64<br>[0.61,0.66] |
|  | Anti-depressant | 0.75<br>[0.74,0.77] | 0.75<br>[0.74,0.77] | 0.73<br>[0.71,0.75] | 0.71<br>[0.67,0.73] |
|  | Anti-psychotic | 0.79<br>[0.75,0.81] | 0.78<br>[0.74,0.81] | 0.76<br>[0.71,0.80] | 0.76<br>[0.73,0.78] |
| ABCD | Stimulant | 0.70<br>[0.70,0.71] | 0.69<br>[0.68,0.70] | 0.69<br>[0.67,0.70] | 0.66<br>[0.65,0.67] |
|  | Anti-depressant | 0.67<br>[0.65,0.69] | 0.66<br>[0.63,0.69] | 0.67<br>[0.64,0.69] | 0.65<br>[0.61,0.67] |
|  | Anti-psychotic | 0.81<br>[0.79,0.83] | 0.80<br>[0.77,0.82] | 0.82<br>[0.78,0.84] | 0.77<br>[0.74,0.80] |
| PPP | Stimulant | 0.84<br>[0.81,0.88] | 0.84<br>[0.80,0.88] | 0.80<br>[0.76,0.84] | 0.78<br>[0.73,0.81] |
|  | Anti-depressant | 0.82<br>[0.77,0.87] | 0.83<br>[0.76,0.88] | 0.79<br>[0.74,0.82] | 0.75<br>[0.71,0.80] |
|  | Anti-psychotic | 0.87<br>[0.83,0.90] | 0.88<br>[0.84,0.90] | 0.84<br>[0.76,0.86] | 0.81<br>[0.76,0.86] |

PPP: Psychopharmacology Program; POND: Province of Ontario Neurodevelopmental network; HBN: Healthy Brain Network; ABCD: Adolescent Brain Cognitive Development study

#### Non-stimulants

Models were trained and evaluated for non-stimulant prescribing in the research and EMR cohorts using the same methodology as the primary medication classes; AU-ROC, AU-PR, sensitivity, and specificity are reported (Supplemental Table 6). This class was not included in the main analysis due to variability in its clinical classification.

**Supplemental Table 6.**
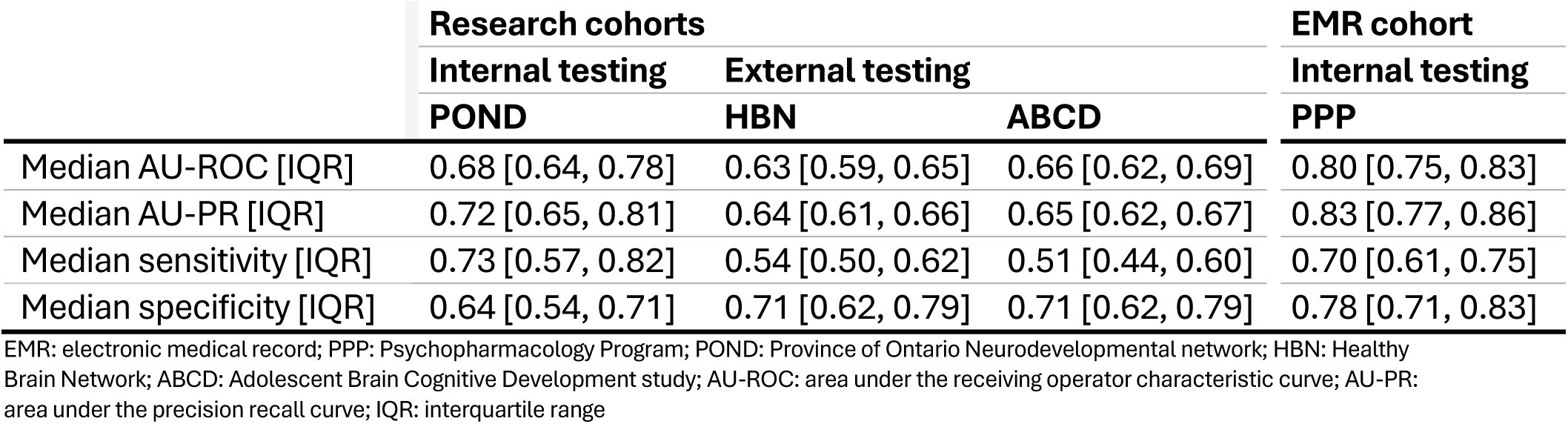
Non-stimulant model performance.

|  | Research cohorts |  |  | EMR cohort |
| --- | --- | --- | --- | --- |
|  | Internal testing | External testing |  | Internal testing |
|  | POND | HBN | ABCD | PPP |
| Median AU-ROC [IQR] | 0.68 [0.64, 0.78] | 0.63 [0.59, 0.65] | 0.66 [0.62, 0.69] | 0.80 [0.75, 0.83] |
| Median AU-PR [IQR] | 0.72 [0.65, 0.81] | 0.64 [0.61, 0.66] | 0.65 [0.62, 0.67] | 0.83 [0.77, 0.86] |
| Median sensitivity [IQR] | 0.73 [0.57, 0.82] | 0.54 [0.50, 0.62] | 0.51 [0.44, 0.60] | 0.70 [0.61, 0.75] |
| Median specificity [IQR] | 0.64 [0.54, 0.71] | 0.71 [0.62, 0.79] | 0.71 [0.62, 0.79] | 0.78 [0.71, 0.83] |

EMR: electronic medical record; PPP: Psychopharmacology Program; POND: Province of Ontario Neurodevelopmental network; HBN: Healthy Brain Network; ABCD: Adolescent Brain Cognitive Development study; AU-ROC: area under the receiving operator characteristic curve; AU-PR: area under the precision recall curve; IQR: interquartile range

#### Excluding prior use

To assess the extent to which PPP model performance depended on prior medication use as the first visit, we re-fit the model after removing the prior use predictors. As expected, given that PPP serves children who have already tried at least one psychoactive medication, performance decreased across all three medication classes (Supplemental Table 7). However, all models remained above chance, indicating that the remaining demographic, clinical, family-related, child-related, and care-related features carry independent predictive signal.

**Supplemental Table 7.** Median [IQR] AU-ROC for the full EMR model compared to when prior medication usage is excluded as a predictor.

|  | Full model | Excluding prior use |
| --- | --- | --- |
| Stimulant | 0.84 [0.81, 0.88] | 0.75 [0.71, 0.78] |
| Anti-depressant | 0.82 [0.77, 0.87] | 0.72 [0.68, 0.78] |
| Anti-psychotic | 0.87 [0.83, 0.91] | 0.66 [0.61, 0.70] |

AU-ROC: area under the receiver operating characteristic curve; IQR: interquartile range; EMR: electronic medical record

#### Per-group fairness rates

The per-group rates underlying the DPR and EOR ratios reported in Table 2 for the research and EMR cohorts are provided in Supplemental Tables 8 and 9, respectively, including the percentage of participants predicted positive (PP; underlying DPR), the true and false positive rates (TPR, FPR; underlying EOR), and classification accuracy for each sensitive group.

**Supplemental Table 8.**
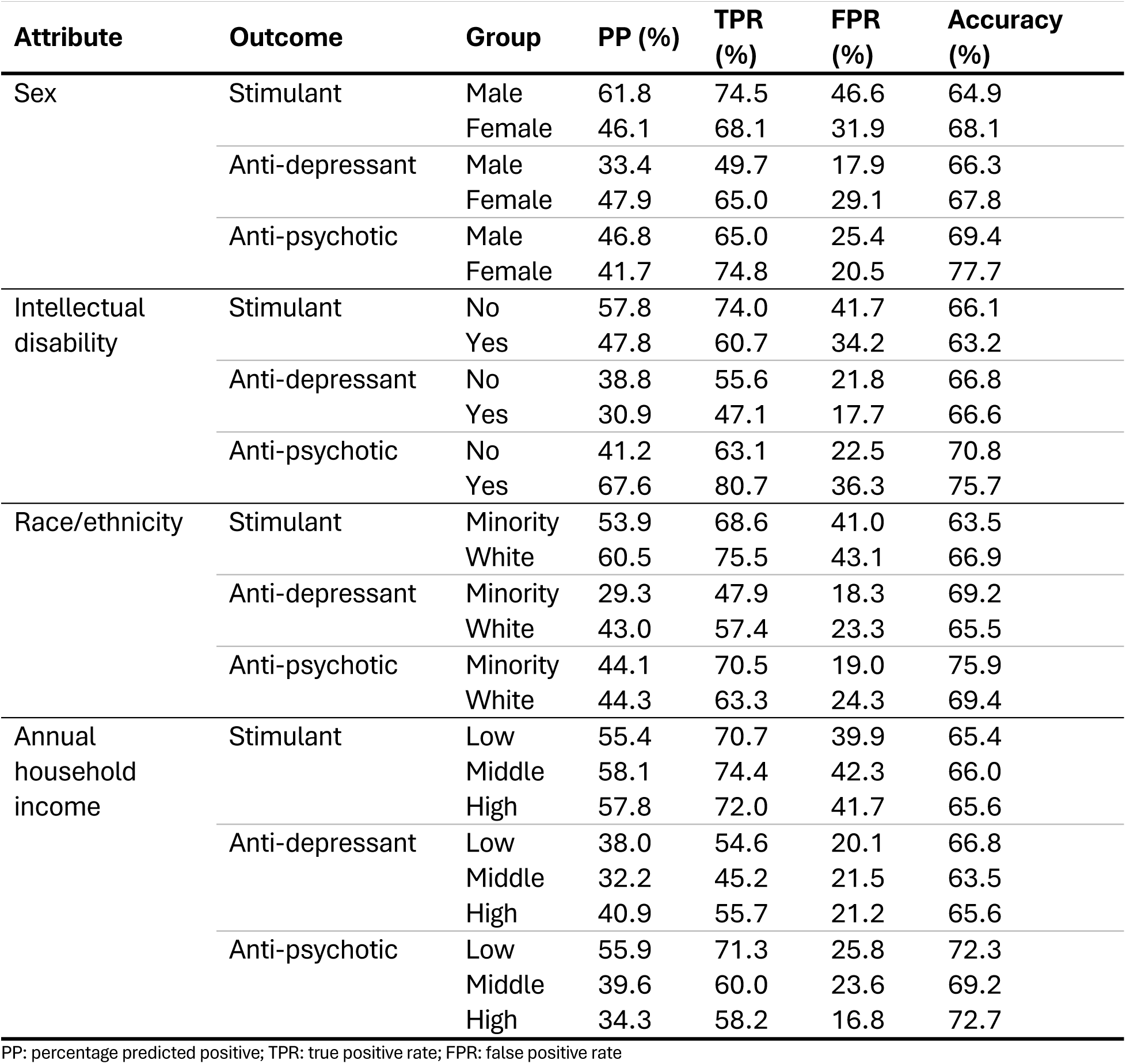
Per-group rates underlying the demographic parity (DPR) and equalized odds (EOR) ratios for the research cohorts.

**Supplemental Table 9.**
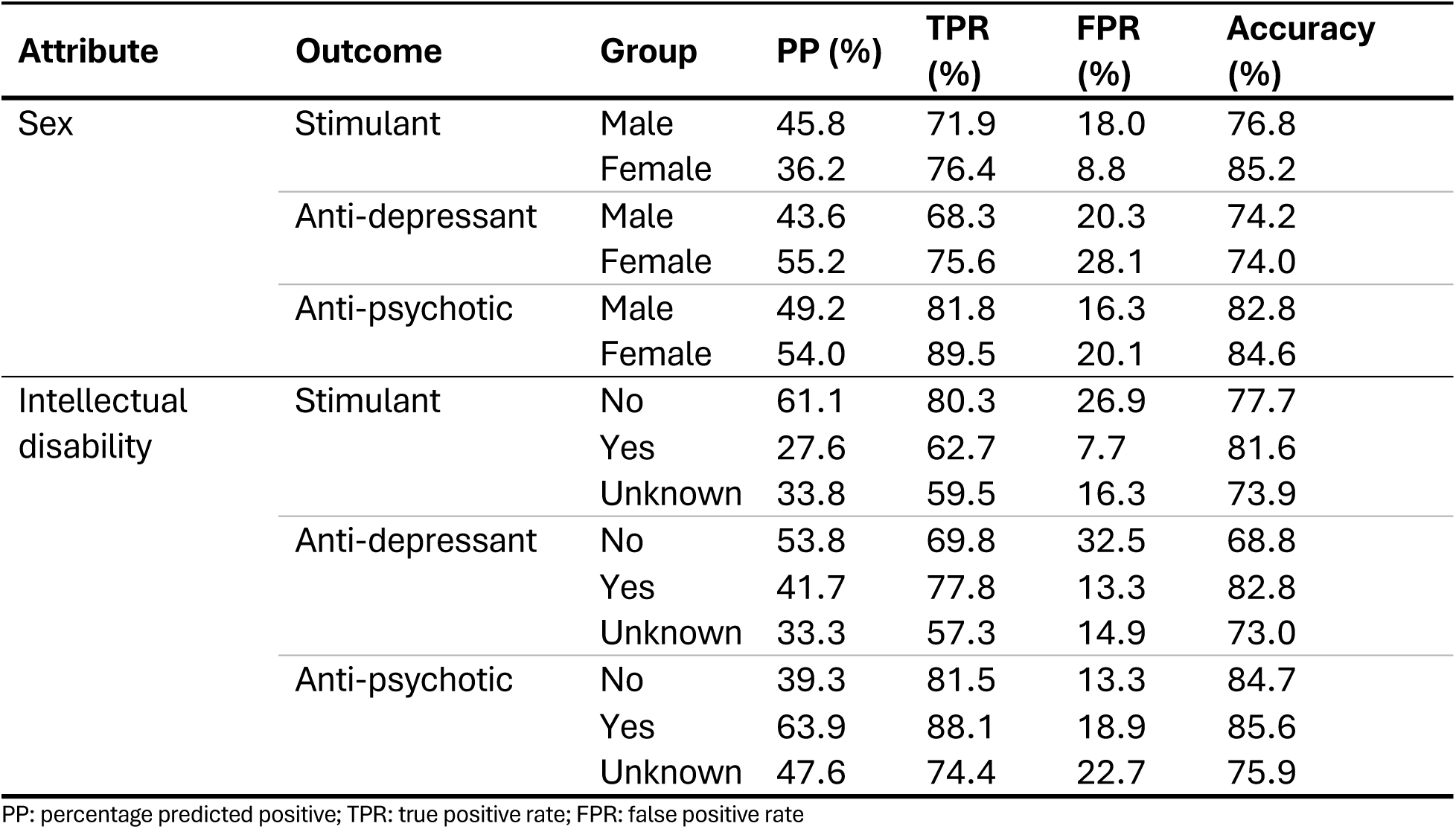
Per-group rates underlying the demographic parity (DPR) and equalized odds (EOR) ratios for the EMR cohort.

| Attribute | Outcome | Group | PP (%) | TPR (%) | FPR (%) | Accuracy (%) |
| --- | --- | --- | --- | --- | --- | --- |
| Sex | Stimulant | Male | 45.8 | 71.9 | 18.0 | 76.8 |
|  |  | Female | 36.2 | 76.4 | 8.8 | 85.2 |
|  | Anti-depressant | Male | 43.6 | 68.3 | 20.3 | 74.2 |
|  |  | Female | 55.2 | 75.6 | 28.1 | 74.0 |
|  | Anti-psychotic | Male | 49.2 | 81.8 | 16.3 | 82.8 |
|  |  | Female | 54.0 | 89.5 | 20.1 | 84.6 |
| Intellectual disability | Stimulant | No | 61.1 | 80.3 | 26.9 | 77.7 |
|  |  | Yes | 27.6 | 62.7 | 7.7 | 81.6 |
|  |  | Unknown | 33.8 | 59.5 | 16.3 | 73.9 |
|  | Anti-depressant | No | 53.8 | 69.8 | 32.5 | 68.8 |
|  |  | Yes | 41.7 | 77.8 | 13.3 | 82.8 |
|  |  | Unknown | 33.3 | 57.3 | 14.9 | 73.0 |
|  | Anti-psychotic | No | 39.3 | 81.5 | 13.3 | 84.7 |
|  |  | Yes | 63.9 | 88.1 | 18.9 | 85.6 |
|  |  | Unknown | 47.6 | 74.4 | 22.7 | 75.9 |

PP: percentage predicted positive; TPR: true positive rate; FPR: false positive rate

#### Baseline demographic parity ratio

The baseline demographic parity ratio (DPR) was computed from actual prescribing labels in each cohort (Supplemental Table 10) to serve as a reference for the model-predicted DPR (Table 2), allowing assessment of whether the model attenuates or amplifies existing prescribing disparities.

**Supplemental Table 10.**
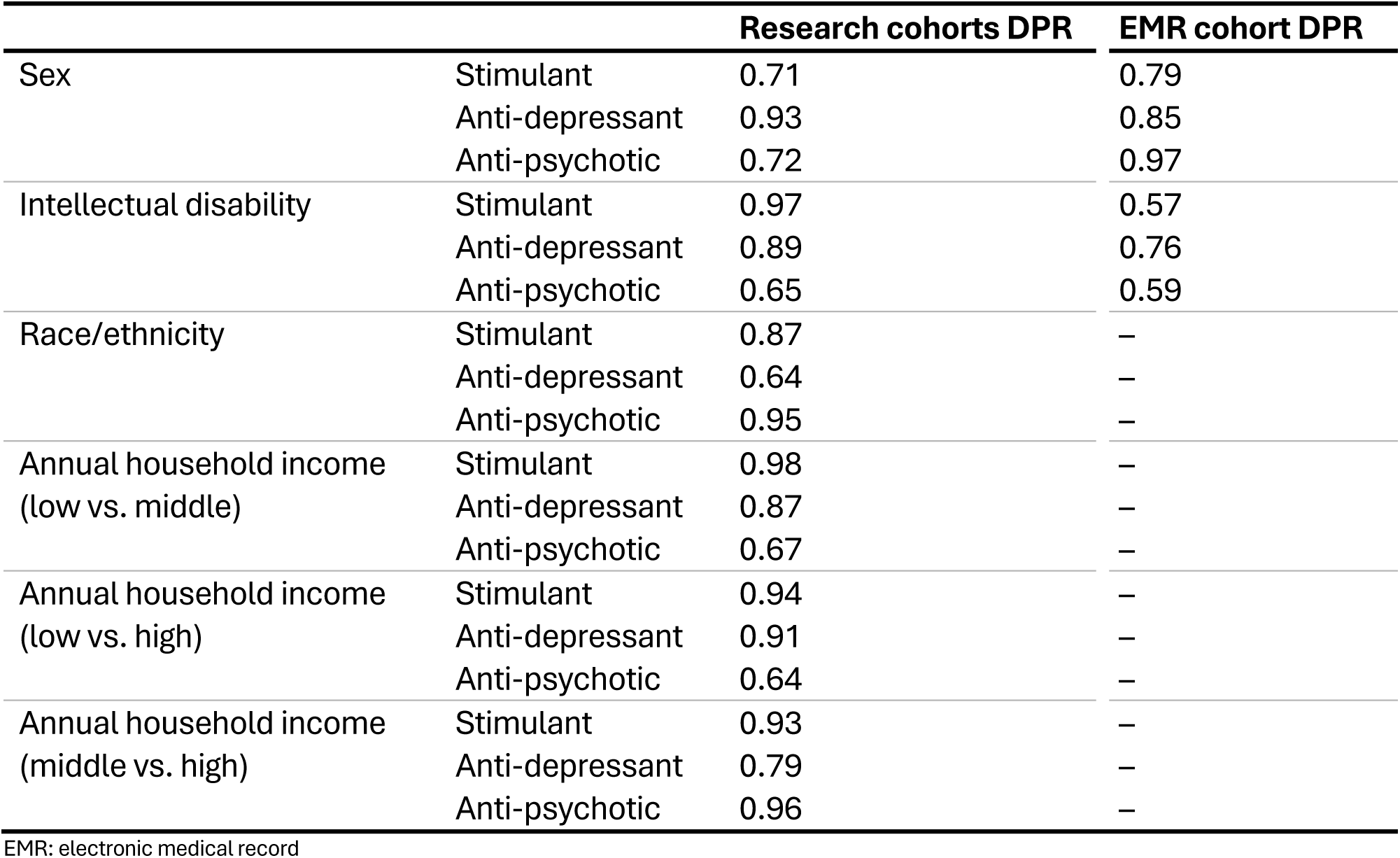
Baseline demographic parity ratio (DPR) computed from actual prescribing labels for sensitive groups.

## Data Availability

Data from four cohorts were used in this study, each governed by their own data sharing restrictions; full details can be found in the Supplementary Material. Code used in this manuscript is available at GitHub (https://github.com/marlvan/med_prediction).

## Acknowledgements

Funding was provided by the Canadian Institutes of Health Research (Operating Grant #527447). This research was undertaken, in part, thanks to funding from the Canada Research Chairs Program. Acknowledgements for each of the research cohorts (POND, HBN, and ABCD) used in this study are provided below.

## POND

This research was conducted with the support of the Ontario Brain Institute (POND, PIs: Dr. Evdokia Anagnostou and Dr. Jason P. Lerch), an independent non-profit corporation, funded partially by the Ontario government. The opinions, results and conclusions are those of the authors and no endorsement by the Ontario Brain Institute is intended or should be inferred.

## HBN

This manuscript was prepared using a limited access dataset obtained from the Child Mind Institute Biobank, the Healthy Brain Network (HBN). This manuscript reflects the views of the authors and does not necessarily reflect the opinions or views of the Child Mind Institute.

## ABCD

Data used in the preparation of this article were obtained from the Adolescent Brain Cognitive DevelopmentSM (ABCD) Study (https://abcdstudy.org), held in the NIMH Data Archive (NDA). This is a multisite, longitudinal study designed to recruit more than 10,000 children age 9-10 and follow them over 10 years into early adulthood. The ABCD Study® is supported by the National Institutes of Health and additional federal partners under award numbers U01DA041048, U01DA050989, U01DA051016, U01DA041022, U01DA051018, U01DA051037, U01DA050987, U01DA041174, U01DA041106, U01DA041117, U01DA041028, U01DA041134, U01DA050988, U01DA051039, U01DA041156, U01DA041025, U01DA041120, U01DA051038, U01DA041148, U01DA041093, U01DA041089, U24DA041123, U24DA041147. A full list of supporters is available at https://abcdstudy.org/federal-partners.html. A listing of participating sites and a complete listing of the study investigators can be found at https://abcdstudy.org/consortium_members/. ABCD consortium investigators designed and implemented the study and/or provided data but did not necessarily participate in the analysis or writing of this report. This manuscript reflects the views of the authors and may not reflect the opinions or views of the NIH or ABCD consortium investigators. The ABCD data repository grows and changes over time. The ABCD data used in this report came from DOI: 10.15154/z563-zd24. DOIs can be found at https://nda.nih.gov/abcd

